# Awareness and Agreement with Neurofibromatosis Care Guidelines among Neurofibromatosis Specialists

**DOI:** 10.1101/2021.06.16.21259047

**Authors:** Vanessa L. Merker, Pamela Knight, Heather B. Radtke, Kaleb Yohay, Nicole J. Ullrich, Scott R. Plotkin, Justin T. Jordan

**Affiliations:** Department of Neurology and Cancer Center, Massachusetts General Hospital, Boston, MA USA, 02144; Center for Healthcare Organization and Implementation Research (CHOIR), VA Bedford Healthcare System, Bedford, MA, USA 01730; Children’s Tumor Foundation, New York, New York, USA, 10017; Division of Genetics, Medical College of Wisconsin, Milwaukee, Wisconsin, USA, 53226; Department of Neurology, NYU Langone Health, New York, New York, USA, 10017; Department of Neurology, Boston Children’s Hospital, Boston, Massachusetts, USA, 02115

**Keywords:** neurofibromatosis 1, neurofibromatosis 2, schwannomatosis, practice guidelines

## Abstract

**Introduction:** Given the wide range of neurofibromatoses (NF) symptoms and medical specialties involved in NF care, we sought to evaluate the level of awareness of, and agreement with, published NF clinical guidelines among United States NF specialists.

**Methods:** An anonymous, cross-sectional online survey was distributed to attendees of a large NF research conference. Respondents self-reported demographics, practice characteristics, awareness of seven NF guideline publications, and level of agreement with up to 40 individual guidelines using a 5-point Likert scale. We calculated the proportion of guidelines that each clinician rated “strongly agree”, and assessed for differences in guideline awareness and agreement by respondent characteristics.

**Results:** Sixty-three clinicians (49% female; 80% academic practice) across >8 medical specialties completed the survey. Awareness of each guideline publication ranged from 53%-79% of respondents; specialists had higher awareness of publications endorsed by their medical professional organization (p<0.05). The proportion of respondents who “strongly agree” with individual guidelines ranged from 17%-83%; for 16 guidelines, less than 50% of respondents “strongly agree”. There were no significant differences in overall agreement with guidelines based on clinicians’ gender, race, specialty, years in practice, practice type (academic/private practice/other), practice location (urban/suburban/rural), or involvement in NF research (p>0.05 for all).

**Conclusions:** We identified wide variability in both awareness of, and agreement with, published NF care guidelines among NF experts. Future efforts should focus on evidence-based, consensus-driven methods to update and disseminate guidelines across this multi-specialty group. Patients and caregivers should also be consulted to anticipate barriers to accessing and implementing guideline-driven care.

## Introduction

The neurofibromatoses (NF) - neurofibromatosis 1 (NF1), neurofibromatosis 2 (NF2) and schwannomatosis - are rare genetic disorders that predispose patients to develop multiple nerve sheath tumors and many other physical, neurocognitive, and psychosocial symptoms.^1^ Given the varied clinical spectrum of these diseases, physicians from multiple medical specialties provide care for NF patients. For example, patients with NF1 may visit pediatricians for initial evaluations; neurologists or neuro-oncologists for brain tumors, learning disabilities, headaches, or focal neurological deficits; plastic surgeons or dermatologists for cutaneous neurofibromas; orthopedic surgeons for scoliosis or pseudoarthrosis; and various other specialists for their complex disease. The need for specialty care for the wide range of manifestations of NF may lead to fragmented care, highlighting the critical importance of disseminating clinical best practices.

Clinical practice guideline publications provide clinicians with collated recommendations based on systematic evidence reviews, while expert review documents provide recommendations where evidence-based data is more limited.^2^ Currently there are seven peer-reviewed publications providing clinical care recommendations for NF, hereafter referred to collectively as ‘NF guidelines’.^3-9^ Prior work has shown the importance of clinicians’ knowledge and attitudes - including their familiarity and agreement with guidelines – in predicting implementation of clinical guidelines.^10,11^ However, to date, there is no evidence of how broadly NF guidelines have been disseminated across the various specialties caring for NF patients, nor is there data on the agreement with or use of these guidelines. Therefore, we sought to evaluate the awareness of, and agreement with, NF guidelines among NF clinicians as a first step to developing consensus on NF clinical best practices.

## Materials and Methods

### Participant Recruitment

United States based clinicians (including physicians, advanced practice providers, and other health care providers) who currently care for neurofibromatosis patients were eligible to participate in the survey. Potentially eligible participants were identified using registration lists for the 2016-2018 Children’s Tumor Foundation NF conferences, the largest NF-specific research conference in the U.S. Registration lists did not reliably differentiate whether conference attendees were clinicians or non-clinicians nor their country of residence, so all potentially eligible participants (n=358) were included in the recruitment process and we relied on screening questions at the start of the survey to filter out non-clinicians and those residing outside the U.S. The chair of the Children’s Tumor Foundation Clinical Care Advisory Board emailed survey invitations in September 2019, with a follow-up reminder email two weeks later. Participation in the survey was voluntary and anonymous. The study was deemed exempt by the Mass General Brigham Institutional Review Board.

### Survey Design

This cross-sectional survey was administered online using REDCap, a secure online data collection platform.^12^ Respondents were asked to self-report demographic data and practice characteristics [i.e. gender, race, ethnicity, medical specialty, years in practice, practice type (academic practice/private practice/other), practice location (urban/suburban/rural), involvement in NF research, affiliation with the Children’s Tumor Foundation NF Clinic Network, and whether their practice currently included pediatric and adult NF1, NF2, and schwannomatosis patients.] The research team identified six relevant NF guideline publications^3-8^ via literature review to include in the survey (as French guidelines from Bergqvist et al. (2020) were published after this survey was administered). All respondents were asked to identify whether they were previously aware, unaware, or unsure about each publication [which were identified by first author, title, journal, publication year, PubMed ID and primary topic area (i.e. pediatric NF1)]. Respondents were asked about their awareness of all six NF guideline publications, even if their practice did not currently include the specific NF patient population addressed by the publication, under the assumption that clinicians should have broad knowledge of these resources to remain current with the field, to be prepared to coordinate care for new patients, and to help pediatric patients transition to adult care.

One author (JTJ) extracted 40 individual guidelines from five guideline publications^4-8^ for assessment of guideline agreement (as pediatric NF1 guidelines from Miller et al. (2019) were published after this section of the survey was finalized). Guidelines were largely extracted verbatim, with minor changes in wording to combine nearly identical guidelines from multiple publications or to improve grammar (see Table 1 for full list of guidelines). All guidelines were assessed by two authors (JTJ and VLM) to determine the relevant patient population addressed (i.e. pediatric or adult; NF1, NF2, or schwannomatosis). Respondents were asked to rate their agreement with each individual guideline relevant to their practice population using a 5-point Likert scale (strongly agree to strongly disagree, with neutral as the midpoint). Respondents could also optionally provide free-text comments about any guideline. The study size was determined by the number of eligible respondents who completed at least one guideline agreement rating.

**Table 1.**
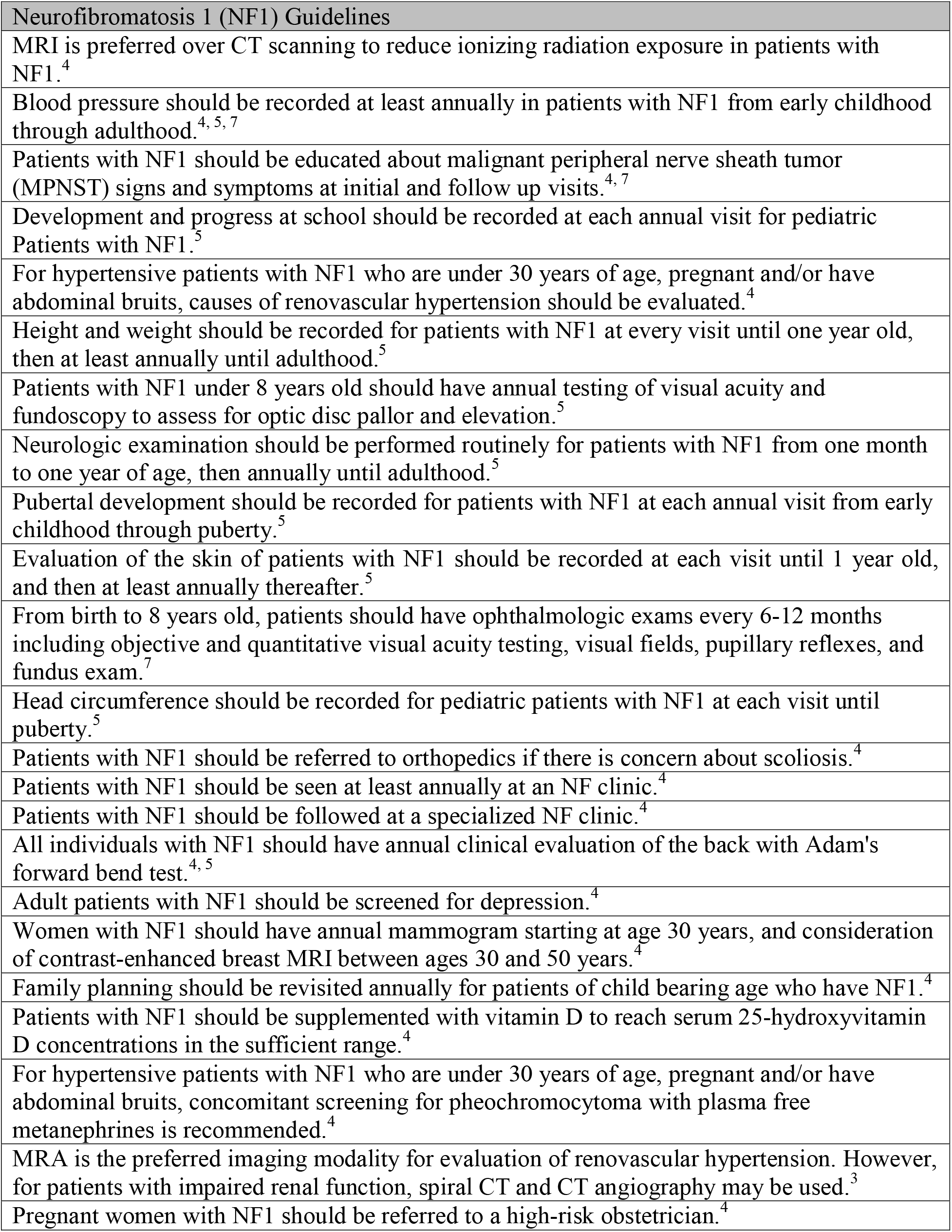
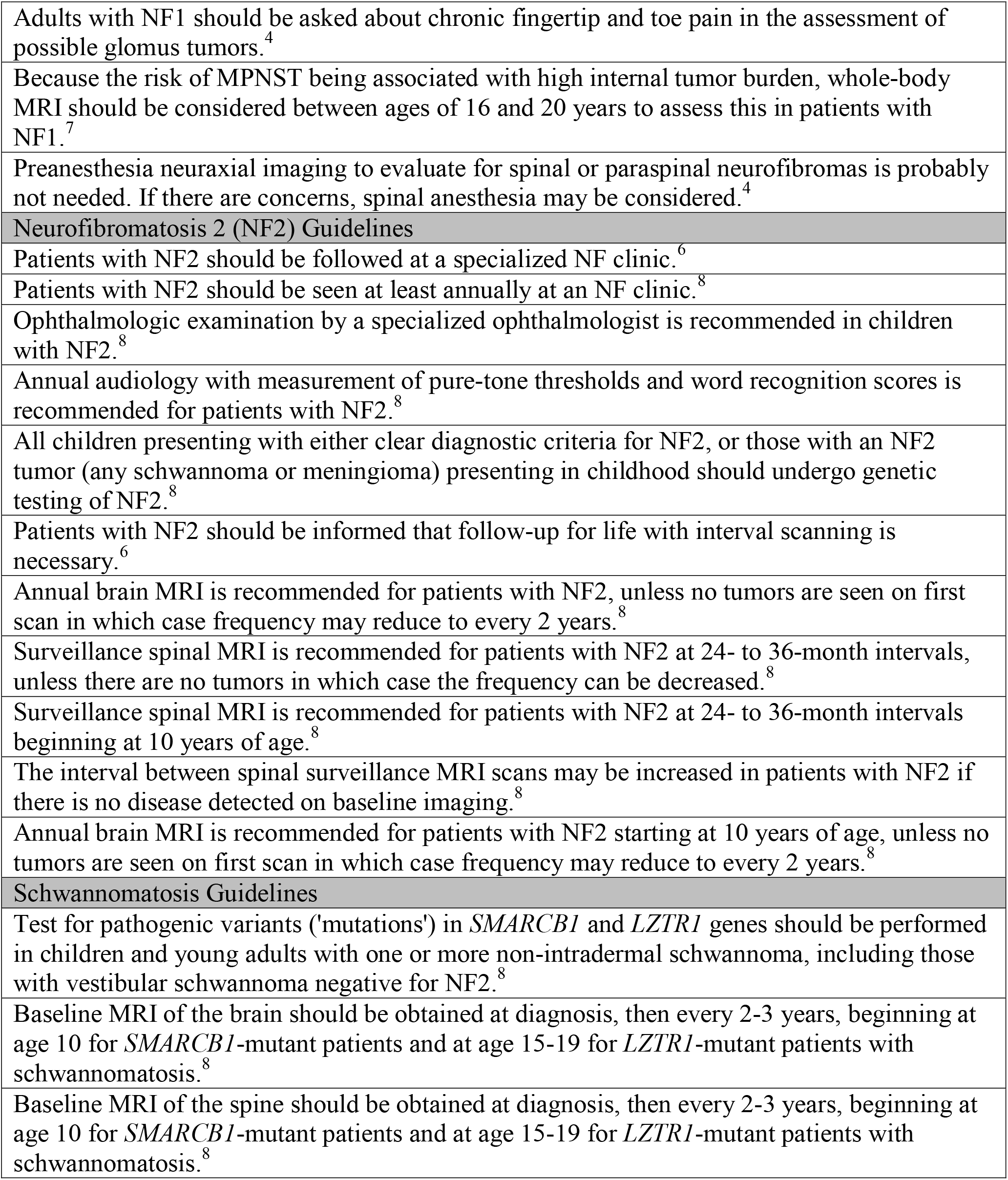
NF Clinical Guidelines Assessed in the Current Study.

### Data Analysis

Descriptive data on all variables are reported as frequencies and percentages. Only questions assessing study eligibility and determining the subpopulations of NF included in each clinicians’ practice were mandatory; all other questions were optional, and percentages are reported out of the sample size of non-missing responses unless otherwise noted. Due to the risk of acquiescence bias (the tendency for survey respondents to select positive response options)^13^, we focused our reporting of guideline agreement on the percentage of respondents who selected “strongly agree”, thus actively indicating a strong preference in favor of each guideline. As clinicians’ level of support for a clinical practice guideline is associated with their adherence to guidelines is practice^14,15^, this type of top-box analysis (used often in patient experience surveys)^16-18^ may best predict clinicians’ real-world behavior. We calculated the 95% confidence interval for the percentage of respondents who strongly agreed with each guideline using standard formulas for margin of error of a proportion; average margin of error across all guidelines was 12.7% (Supplementary Information).

We examined whether clinician demographics or practice characteristics were correlated with level of agreement with NF clinical guidelines. Given the large number of individual guidelines and respondent subgroups of interest (and concomitant risk of Type 1 error from performing multiple comparisons), we assessed clinicians’ overall agreement with all clinical guidelines relevant to their own practice populations. To do this, we calculated the proportion of guidelines to which each clinician ‘strongly agreed’ as a proportion of the total number of guidelines for which they completed ratings. This proportion could range from 0 (does not strongly agree with any guidelines) to 1 (strongly agrees with all guidelines). This proportion was compared across respondent subgroups used t-tests or ANOVA as appropriate.

Two NF1 guidelines publications were endorsed by U.S. medical societies – Miller et al. guidelines for pediatric NF1 published by the American Academy of Pediatrics (AAP) and endorsed by the American College of Medical Genetics and Genomics^3^ (ACMG) and Stewart et al. guidelines for adult NF1 published by ACMG^4^. To better understand guideline dissemination patterns, we assessed whether clinicians in these professions (pediatrics and medical genetics, respectively) were more likely to report being aware of guideline publications than clinicians in other specialties using Fisher’s exact test. A p-value of ≤0.05 was considered statistically significant for all tests. Finally, we used qualitative content analysis to summarize respondent’s free-text comments.^19^

## Results

Eighty-two U.S. based clinicians accessed the survey; twelve did not complete any questions beyond eligibility screening and an additional seven did not complete any guideline agreement questions, resulting in a final analytic sample of 63 people [63/358 (17.6%) of potentially eligible respondents, 63/82 (76.8%) of eligible respondents who accessed the survey]. Clinician demographics, practice characteristics, and type of NF specialization are shown in Table 2. Clinicians represented >8 medical or surgical specialties, with one-third (n=21) of respondents in neurology or neuro-oncology. Clinicians were located across the U.S., representing 26 U.S. states with the greatest number of respondents from New York, California, Florida, Massachusetts, and Minnesota (respectively). Most respondents (n=50, 79.4%) provided clinical care as part of a specialized neurofibromatosis clinic affiliated with the Children’s Tumor Foundation NF Clinic Network^20^ and the majority were involved in NF research (n=39 for clinical trials and n=47 for non-treatment research studies such as tumor banks, natural history studies or questionnaire-based research). These results are similar to the geographic distribution and proportion of NF Clinic Network clinics who were involved in clinical trials at this time [31/56 (55.4%) of clinics compared to 61.9% of survey respondents, per Children’s Tumor Foundation internal 2019 annual report].

**Table 2.** Respondent Demographics and Practice Characteristics (n=63)

| Clinician Demographics | N (%) |
| --- | --- |
| Gender |  |
| Female | 31 (49.2%) |
| Male | 27 (42.9%) |
| Non-binary | 1 (1.6%) |
| Prefer not to answer or missing | 4 (6.3%) |
| Race |  |
| White | 53 (84.1%) |
| Black or African American | 1 (1.6%) |
| Asian | 6 (8.4%) |
| American Indian or Alaska Native | 0 (0.0%) |
| Native Hawaiian or other Pacific Islander | 0 (0.0%) |
| Other | 2 (3.2%) |
| Missing | 1 (1.6%) |
| Ethnicity |  |
| Hispanic or Latino | 3 (4.8%) |
| Not Hispanic or Latino | 56 (88.9%) |
| Missing | 4 (6.3%) |
| Primary Specialty |  |
| Medical Genetics | 18 (28.6%) |
| Neuro-Oncology | 11 (17.4%) |
| Neurology | 10 (15.9%) |
| Pediatrics | 5 (7.9%) |
| Hematology/Oncology or Medical Oncology | 5 (7.9%) |
| Dermatology | 2 (3.2%) |
| Neurosurgery | 1 (1.6%) |
| Orthopedic Surgery | 0 (0.0%) |
| Other | 8 (13.7%) |
| Missing | 3 (4.8%) |
| Years in Post-Training Practice |  |
| <5 years | 11 (17.4%) |
| 5-10 years | 14 (22.2%) |
| 10-20 years | 21 (33.3%) |
| >20 years | 17 (27.0%) |
| Practice Characteristics | N (%) |
| Practice Type |  |
| Academic medical practice | 50 (79.4%) |
| Private Practice | 5 (7.9%) |
| Other | 7 (11.1%) |
| Missing | 1 (1.6%) |
| Location of Primary Office |  |
| Urban | 49 (77.8%) |
| Suburban | 12 (19.0%) |
| Rural | 1 (1.6%) |
| Missing | 1 (1.6%) |
| Primary Language of Patients in Practice |  |
| English | 61 (96.8%) |
| Spanish | 2 (3.2%) |
| Neurofibromatosis Specialization | N (%) |
| Affiliation with Children's Tumor Foundation<br>NF Clinic Network |  |
| Yes | 50 (79.4%) |
| No | 11 (17.4%) |
| Unsure | 1 (1.6%) |
| Missing | 1 (1.6%) |
| Clinician Involvement in NF Treatment Trials |  |
| Yes | 39 (61.9%) |
| No | 23 (36.5%) |
| Missing | 1 (1.6%) |
| Clinician Involvement in NF Non-Treatment<br>Research |  |
| Yes | 47 (74.6%) |
| No | 16 (25.4%) |
| Clinicians Seeing Each Patient Population |  |
| Pediatric Neurofibromatosis 1 | 55 (87.3%) |
| Adult Neurofibromatosis 1 | 51 (81.0%) |
| Pediatric Neurofibromatosis 2 | 48 (76.2%) |
| Adult Neurofibromatosis 2 | 36 (57.1%) |
| Schwannomatosis | 36 (57.1%) |

### Guideline Awareness

Clinicians self-reported their awareness of publications containing NF clinical care guidelines (Table 3). The percentage of respondents who were aware of each guideline set ranged from 53.2% (for schwannomatosis guidelines within Evans et al. 2017) to 79.4% (for pediatric NF1 guidelines published by Ferner et al. 2007). Among only those respondents who reported currently seeing the relevant patient population, awareness was only marginally increased (by 1.1 to 5.1 percentage points), with the exception of adult NF2 guidelines by Evans et al. 2005, in which awareness increased 11.6 percentage points to 65.7% of respondents. Overall, 26-36% of respondents were unaware of recently published guideline documents (2017-2019). Medical geneticists were significantly more likely to report awareness of adult NF1 guidelines endorsed by the ACMG than clinicians of other specialties (100% of medical geneticists vs. 56.1% of other specialists, p=0.0008). Pediatricians and medical geneticists were also more likely to report awareness of pediatric NF1 guidelines endorsed by the AAP and the ACMG (87.0% of pediatricians and medical geneticists vs. 55.6% of other specialists, p=0.021).

**Table 3.** Clinicians’ Self-Reported Awareness of NF Clinical Guideline Publications.

|  | Total<br>Number of<br>Respondents<br>(N) | Aware of<br>Guideline<br>Document<br>N (%) | Unaware of<br>Guideline<br>Document<br>N (%) | Unsure<br>N (%) |
| --- | --- | --- | --- | --- |
| **Neurofibromatosis 1 Guidelines** |  |  |  |  |
| Stewart et al. (2018) | 62 | 43 (69.4%) | 16 (25.8%) | 3 (4.8%) |
| Ferner et al. (2007) | 63 | 50 (79.4%) | 11 (17.5%) | 2 (3.2%) |
| Evans et al. (2017a) | 63 | 47 (58.7%) | 21 (33.3%) | 5 (7.9%) |
| Miller et al. (2019) | 62 | 43 (69.4%) | 17 (27.4%) | 2 (3.2%) |
| **Neurofibromatosis 2 Guidelines** |  |  |  |  |
| Evans et al. (2005) | 61 | 33 (54.1%) | 26 (42.6%) | 2 (3.3%) |
| Evans et al. (2017b)* | 61 | 40 (65.6%) | 18 (29.5%) | 3 (4.9%) |
| **Schwannomatosis Guidelines** |  |  |  |  |
| Evans et al. (2017b)* | 62 | 33 (53.2%) | 22 (35.5%) | 7 (11.3%) |

Table Legend: \*Respondents were asked to rate their awareness of the neurofibromatosis 2 guidelines and schwannomatosis guideline within Evans et al. 2017b separately.

### Guideline Agreement

Overall, less than half of survey respondents had strong agreement with 16/40 (40%) of NF guidelines. Clinicians’ level of agreement with NF1 guidelines are presented in Figure 1; number of respondents and 95% confidence intervals are available in supplementary table 1. Level of strong agreement with individual guidelines ranged from 17% to 83%. Strong agreement was highest for the preference of MRIs over CT scans to reduce exposure to ionizing radiation exposure (83%); annual blood pressure checks (80%); education about signs and symptoms of malignant peripheral nerve sheath tumors (76%); and annual check-ins on development and school progress for pediatric patients (76%). For 9/26 (34.6%) of NF1 guidelines, less than half of respondents selected ‘strongly agree’. These guidelines addressed breast cancer screening, counseling regarding family planning, vitamin D supplementation, evaluation of hypertension, pregnancy management, assessment of glomus tumors, and use of whole-body MRI.

**Figure 1.**
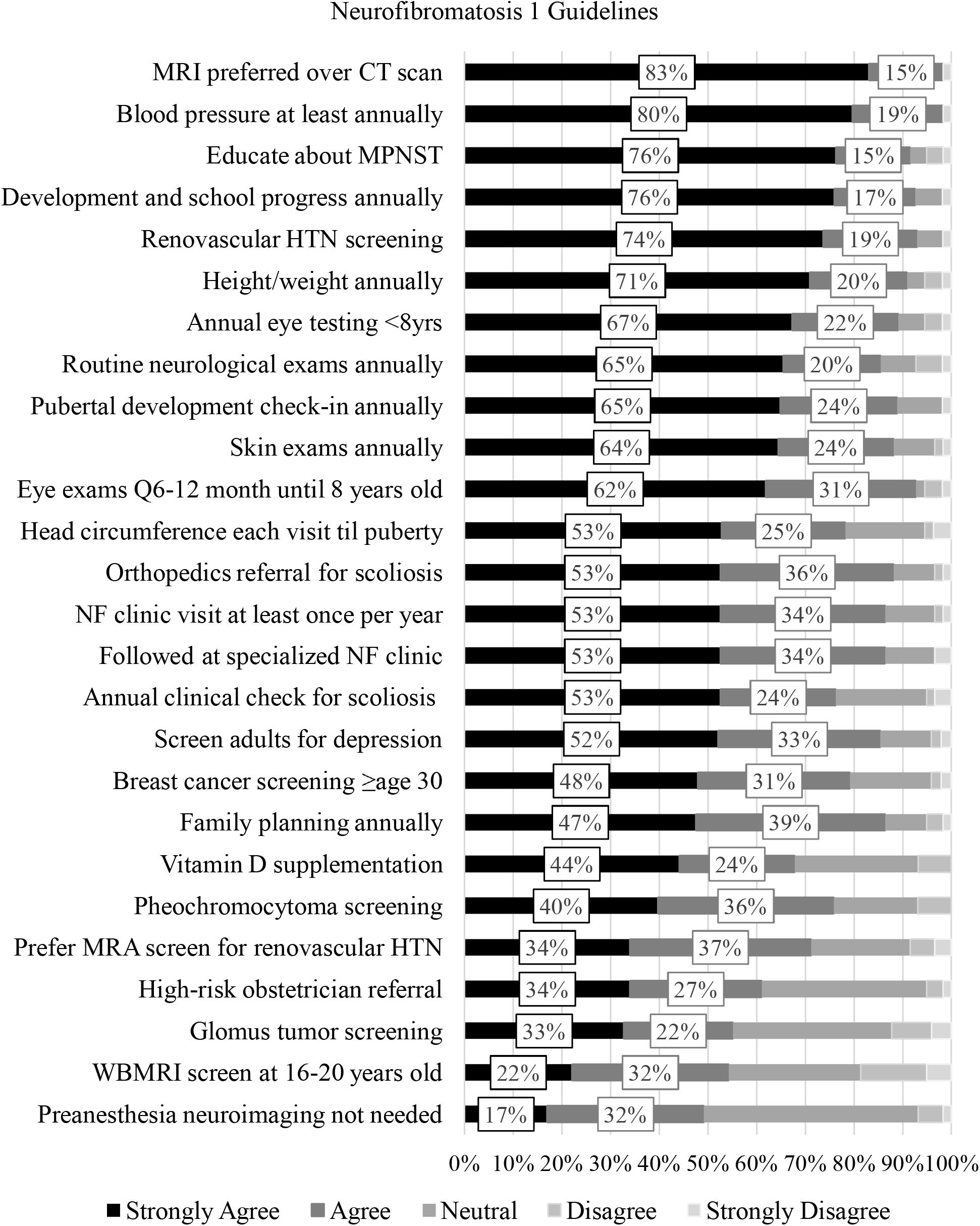
NF Clinician Agreement with Neurofibromatosis 1 Guidelines. Stacked bar charts displaying the percentage of clinicians who “strongly agreed” (dark blue), “agreed” (orange), were “neutral” (gray), “disagreed” (yellow), or “strongly disagreed” (light blue) with each guideline. Overlaid boxes display the percentage of clinicians who “strongly agreed” (dark blue) and “agreed” (orange) to each guideline, rounded to the nearest percentage point. Guidelines are presented with short descriptions for reference; for full guideline text and citations please refer to Table 1, where all guidelines are presented in the same rank order. Abbreviations: CT = computerized tomography; HTN = hypertension; NF = neurofibromatosis; MPNST = malignant peripheral nerve sheath tumor; MRA = magnetic resonance angiography; MRI = magnetic resonance imaging; WBMRI = whole body magnetic resonance imaging.

Clinicians’ level of agreement with NF2 and schwannomatosis guidelines are presented in Figure 2; number of respondents and 95% confidence intervals are available in supplementary table 1. Level of strong agreement with individual NF2 guidelines ranged from 36% to 73%. Strong agreement was highest for receiving care at specialized NF clinics (73%) and for receiving care annually at an NF clinic (72%). For 4/11 (36.4%) of NF2 guidelines, less than half or respondents selected ‘strongly agree’. These guidelines addressed the frequency of spinal MRIs in all patients and the timing of brain MRIs for pediatric patients. Agreement was noticeably lower for pediatric imaging recommendations including the provision to start surveillance at age 10 when compared to identical adult guidelines (absolute difference of 15.4 percentage points for brain MRIs and 9.3 percentage points for spinal MRIs). Level of strong agreement with three schwannomatosis guidelines addressing genetic testing for younger patients with potential schwannomatosis and the timing of brain and spine MRIs (including age at baseline scan and frequency of imaging) ranged from 27-38%.

**Figure 2:**
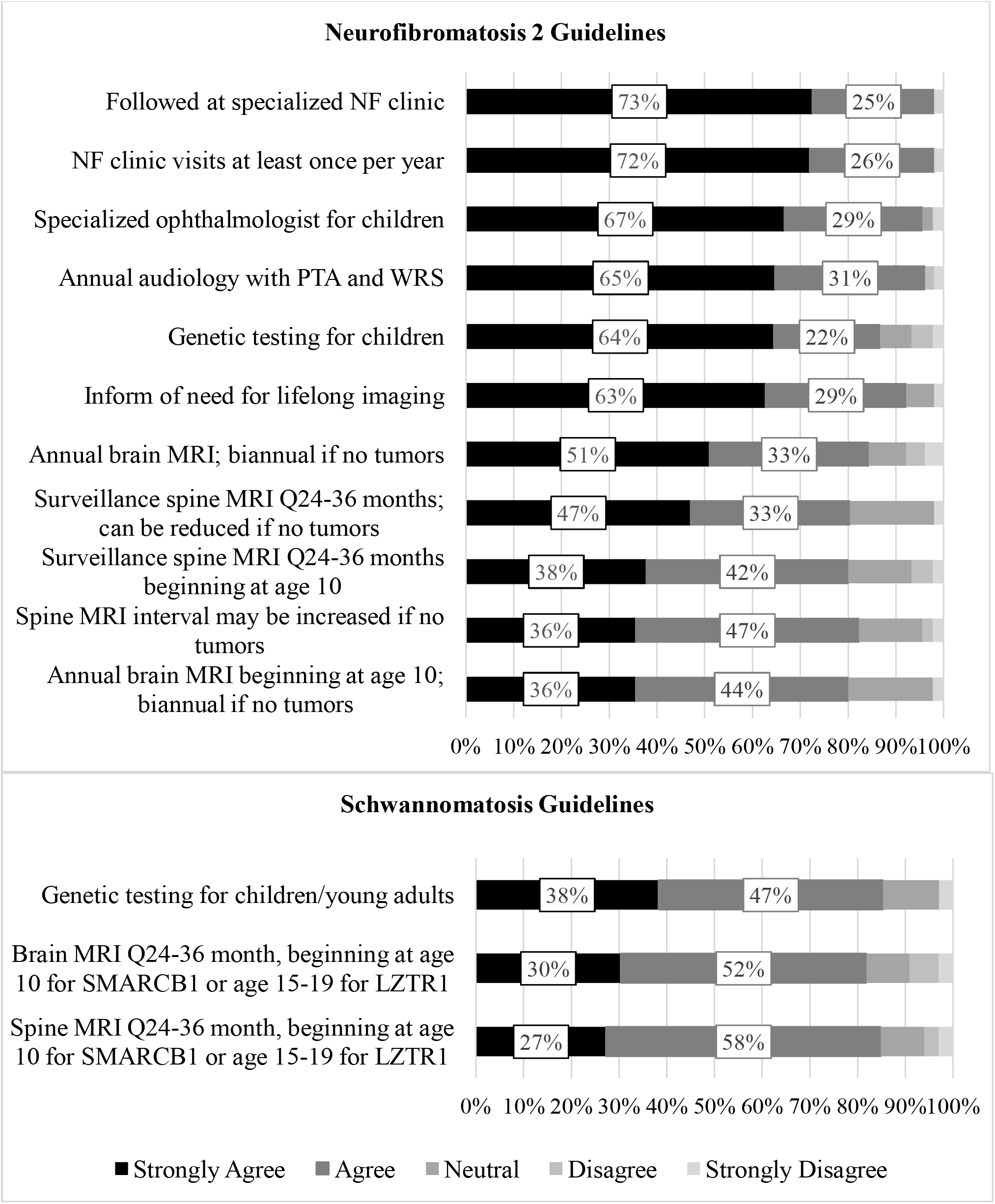
NF Clinician Agreement with Neurofibromatosis 2 and Schwannomatosis Guidelines. Stacked bar charts displaying the percentage of clinicians who “strongly agreed” (dark blue), “agreed” (orange), were “neutral” (gray), “disagreed” (yellow), or “strongly disagreed” (light blue) with each guideline. Overlaid boxes display the percentage of clinicians who “strongly agreed” (dark blue) and “agreed” (orange) to each guideline, rounded to the nearest percentage point. Guidelines are presented with short descriptions for reference; for full guideline text and citations please refer to Table 1, where all guidelines are presented in the same rank order. Abbreviations: NF = neurofibromatosis; MRI = magnetic resonance imaging; PTA = pure tone average; WRS = word recognition score. *SMARCB1* and *LZTR1* refer to gene variants known to cause schwannomatosis.

The median proportion of guidelines with which each clinician strongly agreed was 0.55 (range, 0-1; 25^th^-75^th^ percentile, 0.35 to 0.73). There were no statistically significant differences in guideline agreement proportion based on clinicians’ gender, race, specialty, years in practice, practice type, practice location, participation in the NF Clinic Network, involvement in NF treatment trials, or involvement in non-treatment NF research. Medical geneticists were no more likely to than other specialists to strongly agree with adult NF1 guidelines endorsed by the ACMG (agreement index 0.52 vs. 0.53, p=0.89). As guidelines were not systematically extracted from the pediatric NF1 guidelines endorsed by the AAP and ACMG, a parallel test of pediatricians’ agreement with these guidelines was not performed.

Twenty optional free-text comments regarding NF guidelines were collected from eleven survey respondents. While there were insufficient comments to robustly explain disagreement with individual guidelines, categories of comments may illuminate potential sources of disagreement with NF guidelines in general. Comments were categorized into five main groups, in descending order of frequency: 1) disagreement about the timing of services recommended by a guideline; 2) disagreement that a guideline should apply to the entire population equally; 3) perception that guidelines were not in the provider’s scope of practice; 4) confusion about the way a guideline was worded; and 5) suggested addition to a guideline.

When disagreeing with recommended timing of services, some participants advocated for more frequent monitoring (e.g. “*I think the eye exam with VF/VA [visual fields/visual acuity] should be every 6 months until age 6, and then yearly*” and “*Audiograms are inexpensive and should be done at 6 month intervals*”) while others advocated for less frequent monitoring (e.g., “*I think monthly [neuro] evals would be too frequent. If a baby seems to be doing well at the initial visit, then I see them at 6 months. If I have concerns about development, then I see them at 3 months*.”) When disagreeing with the defined population in a guideline, multiple comments argued that lower risk individuals need less intensive monitoring or referrals (e.g., “*I don’t always refer to Ortho for scoliosis if it is mild and not progressing*” and “*In spite of efforts to standardize imaging interval for spinal imaging, this needs to be individualized, esp. in patients with mild phenotype after adequate surveillance*”) and others argued higher-risk individuals may need more intensive monitoring (e.g. “*If the patients family has an history of aggressive NF2, [starting] imaging earlier than 10 years is recommended*.) Comments regarding scope of practice indicated that not all NF providers treat every NF complication or see a sufficient number of cases to feel prepared to assess the appropriateness of certain guidelines (e.g. *“I am not up to date with NF2 and schwannomatosis, as I do not see adults nor many pediatric patients with these disorders, so I answer “neutral” in those cases”*), and that some guidelines seemed more appropriate for non-specialists to oversee (e.g. “*Some screening such as mammograms are part of routine healthcare*.”) Some comments reflected confusion about the way guidelines were worded (e.g., whether NF1 preanesthesia imaging guidelines were restricted to pregnant individuals and whether NF1 guidelines favoring MRI over CT imaging due to radiation exposure included exceptions for clinical situations where CT may actually be indicated). Finally, one participant suggested that NF1 ophthalmological guidelines include sedated imaging of the brain and orbit if the ophthalmologist has concerns after exam.

## Discussion

We evaluated NF specialists’ agreement with NF clinical guidelines to assess the efficacy of guideline dissemination and identify any areas of disagreement regarding best practices for NF clinical care. We identified wide variability in both awareness of, and agreement with, NF published guidelines among NF experts. While many respondents were aware of relevant guideline publications, and most guidelines did not have large proportions of disagreement, our findings highlight areas warranting further attention as the multidisciplinary NF community strives to reach consensus on optimal care recommendations. Approximately one-quarter to one-third of respondents were unaware of recently published NF guideline documents, and the majority of survey respondents did not strongly agree with 40% of assessed guidelines. While clinician’s primary specialty appears to be related to awareness of NF guidelines, no demographic or practice characteristics were associated with clinicians’ overall level of agreement with NF guidelines.

Clinicians’ awareness of and familiarity with clinical guidelines is the first step in providing guideline-driven care, but can be inhibited by the large volume of information each clinician must process and the limited time they have available to stay informed with literature.^10^ Our findings suggest that clinicians may be more aware of guidelines published by their affiliated medical professional organization (although we could not demonstrate whether they were also more likely to agree with these guidelines). Given the multi-system manifestations of NF and diverse array of specialists involved in NF care, the field should evaluate strategies for broader dissemination of existing guidelines across specialties. Prior research suggests a multifaceted dissemination approach using both written materials (e.g. journal articles, checklists, educational materials, etc.) and personal appeals (e.g. via local discussion forums or one-on-one contacts from peers) may be most effective.^21,22^ Rather than duplicate prior guideline creation efforts, medical societies relevant to NF may also want to review, endorse, and republish relevant guidelines with high agreement to facilitate multidisciplinary collaboration in NF patient care.

However, there remains a number of NF guidelines lacking strong agreement. Potential sources of this reduced enthusiasm for certain guidelines are several-fold. First, perceived lack of evidence on efficacy or utility of a guideline-recommended assessments may limit agreement. For example, the utility of whole body MRI is under study and routine use is increasing; however, the optimal use, timing and interpretation of this technology is not yet known.^4^ Furthermore, not all medical centers have this imaging capability nor do all insurance plans cover whole body MRI for this indication. This inability to reliably implement a guideline-recommended assessment may also lead to lower levels of strong agreement. Another informative example is guidelines on the preferred timing and modality for breast cancer screening in adult women with NF1. There may be lower community agreement on guidelines where additional prospective research evidence is needed (such as comparative effectiveness research on breast MRI vs. mammography) or where standardized guidelines may conflict with individualized patient preferences (such as when personalized shared-decision making is needed to determine optimal screening schedules)^4^ Finally, response to guideline agreement may be impacted by lack of clarity as to which population or clinical context is intended.

This empirical data on agreement with NF clinical guidelines among U.S. specialists can inform discussions on the value of, evidence base for, and implementation of NF clinical guidelines. For guidelines with weaker agreement, consensus-based methods across medical specialties should be used to review guidelines, understand opposition to recommended health services, and propose any necessary clarifications and revisions. Any updated guidelines should adhere to best practices for guideline development.^23^ For example, the Appraisal of Guidelines, Research and Evaluation (AGREE II) instrument can be used to assess the quality of existing clinical guidelines and provide guidance on appropriate development and reporting of revised guidelines.^24^ Many sources emphasize writing clear, unambiguous guidelines that are explicitly and transparently linked to the underlying evidence base.^25,26^ For guidelines with insufficient evidence to support widespread agreement and adoption, funders could prioritize systematic evidence reviews and/or additional prospective studies on the value of recommended interventions.

Recent research also emphasizes the need to incorporate patient perspectives in guideline development.^23,24^ Interdisciplinary teams that include clinicians across multiple specialties and methodologists in implementation science, guideline development, and qualitative research may be best poised to meaningfully incorporate patient preferences into guidelines and address real-world barriers to implementation.^26,27^ It is important to address external barriers such as patients preferences that conflict with guideline recommendations and environmental factors (such as lack of time, resources, or insurance approval) to ensure widespread implementation of guidelines.^10^ Input from clinicians, patients, and family members from diverse practice settings and geographic locations should be sought to understand access and barriers to receiving guideline-driven care. A survey of patients and their family members/caregivers to assess receipt of guideline-concordant care is currently being developed for an NF patient registry for this purpose.^28^

There are several limitations to this study. As we desired to get broad input from the NF community on current guidelines, our recruitment email was sent to a large distribution list and the exact number of eligible clinicians approached and their response rate is unknown. While the exact percentage of NF clinicians who are aware of or agree with guidelines in our cross-sectional sample may vary from the overall population of U.S. NF specialists, our findings highlight important areas of disagreement that likely need to be addressed. Our target population was clinicians attending NF-specific research conferences, who likely specialize in NF and see a high volume of NF patients. This population is likely more aware of NF guideline publications and possibly more likely to agree with guidelines than the larger group of clinicians who care for NF patients. Future work should seek to understand how primary care clinicians find and use guidelines for patients with rare diseases who may be unique in their practice, and how specialists and primary care clinicians can share responsibilities for providing guideline-concordant care for such patients (especially for routine monitoring and surveillance). Finally, due to the large number of guidelines relative to our sample size, we did not statistically test for demographic or practice factors that may influence agreement with individual guidelines. Qualitative work exploring clinicians’ reasons for not strongly agreeing with specific NF guidelines would be informative in revising guidelines and developing additional interventions to promote guideline use.

In conclusion, our study demonstrates that there is significant variation in levels of strong agreement with NF clinical guidelines. For guidelines with strong agreement, future efforts to enhance guideline use should prioritize broad dissemination of guidelines across medical specialties. A focus on implementation of guidelines in routine care – for example, by providing multiple versions of guidelines adapted for different users and purposes, and organizing and formatting content to make it easy to understand and use – will enhance the success of dissemination efforts.^29^ For guidelines with weak or mixed agreement, future efforts to improve guideline relevance and use should prioritize evidence-based, consensus-driven approaches to reviewing and updating guidelines. An international Delphi panel, such as that used recently to revise NF diagnostic criteria, may be an efficient approach to gather widespread input from all relevant stakeholders, including patients and their family members.^30^ Together these initiatives could help combat common barriers to guidelines awareness, agreement, and implementation, maximizing the chances for guideline adherence and the delivery of high-quality NF care in the future.

## Data Availability

While the survey was anonymous, there is a risk of identification from unique combinations of multiple demographic factors in this small group of specialists. For this reason, the full dataset is not publicly available. De-identified data is available upon reasonable request from the corresponding author.

## Declarations

### Code availability

N/A.

### Ethics approval

The study protocol was reviewed by the Mass General Brigham Institutional Review Board and was deemed to meet criteria for exemption.

### Consent to participate

Participation in the survey was voluntary and anonymous. Participants read a short fact sheet about the study and indicated their consent to participate by completing the survey.

### Consent for publication

N/A.

## Acknowledgements

The authors would like to acknowledge the members of the Children’s Tumor Foundation Clinical Care Advisory Board and staff of Children’s Tumor Foundation for helpful feedback on the survey, and express appreciation to the NF clinicians who completed the survey.

## References

1. Plotkin SR, Wick A. Neurofibromatosis and Schwannomatosis. Semin Neurol. 2018; 38(1):73–85.

2. Institute of Medicine (US) Committee on Standards for Developing Trustworthy Clinical Practice Guidelines, Graham R, Mancher M, Miller Wolman D, Greenfield S, Steinberg E. Clinical Practice Guidelines We Can Trust. Washington DC: National Academies Press; 2011: https://www.ncbi.nlm.nih.gov/books/NBK209546/.

3. Miller DT, Freedenberg D, Schorry E, et al. Health Supervision for Children With Neurofibromatosis Type 1. Pediatrics. 2019; 143(5).

4. Stewart DR, Korf BR, Nathanson KL, Stevenson DA, Yohay K. Care of adults with neurofibromatosis type 1: a clinical practice resource of the American College of Medical Genetics and Genomics (ACMG). Genet Med. 2018; 20(7):671–682.

5. Ferner RE, Huson SM, Thomas N, et al. Guidelines for the diagnosis and management of individuals with neurofibromatosis 1. J Med Genet. 2007; 44(2):81–88.

6. Evans DG, Baser ME, O’Reilly B, et al. Management of the patient and family with neurofibromatosis 2: a consensus conference statement. Br J Neurosurg. 2005; 19(1):5–12.

7. Evans DGR, Salvador H, Chang VY, et al. Cancer and Central Nervous System Tumor Surveillance in Pediatric Neurofibromatosis 1. Clin Cancer Res. 2017; 23(12):e46–e53.

8. Evans DGR, Salvador H, Chang VY, et al. Cancer and Central Nervous System Tumor Surveillance in Pediatric Neurofibromatosis 2 and Related Disorders. Clin Cancer Res. 2017; 23(12):e54–e61.

9. Bergqvist C, Servy A, Valeyrie-Allanore L, Ferkal S, Combemale P, Wolkenstein P. Neurofibromatosis 1 French national guidelines based on an extensive literature review since 1966. Orphanet Journal of Rare Diseases. 2020; 15(1).

10. Cabana MD, Rand CS, Powe NR, et al. Why don’t physicians follow clinical practice guidelines? A framework for improvement. JAMA. 1999; 282(15):1458–1465.

11. Fischer F, Lange K, Klose K, Greiner W, Kraemer A. Barriers and Strategies in Guideline Implementation— A Scoping Review. 2016; 4(3):36.

12. Harris PA, Taylor R, Thielke R, Payne J, Gonzalez N, Conde JG. Research electronic data capture (REDCap) - A metadata-driven methodology and workflow process for providing translational research informatics support. J Biomed Inform. 2009; 42(2):377–381.

13. Krosnick JA. Response strategies for coping with the cognitive demands of attitude measures in surveys. Applied Cognitive Psychology. 1991; 5(3):213–236.

14. Francke AL, Smit MC, de Veer AJE, Mistiaen P. Factors influencing the implementation of clinical guidelines for health care professionals: a systematic meta-review. BMC medical informatics and decision making. 2008; 8:38–38.

15. Bierbaum M, Rapport F, Arnolda G, et al. Clinicians’ attitudes and perceived barriers and facilitators to cancer treatment clinical practice guideline adherence: a systematic review of qualitative and quantitative literature. Implementation Science. 2020; 15(1):39.

16. Lapin BR, Honomichl RD, Thompson NR, et al. Association Between Patient Experience With Patient-Reported Outcome Measurements and Overall Satisfaction With Care in Neurology. Value in Health. 2019; 22(5):555–563.

17. Hanson KT, Zalewski NL, Hocker SE, Caselli RJ, Habermann EB, Thiels CA. At the Intersection of Patient Experience Data, Outcomes Research, and Practice: Analysis of HCAHPS Scores in Neurology Patients. Mayo Clinic Proceedings: Innovations, Quality & Outcomes. 2018; 2(2):137–147.

18. Indovina K, Keniston A, Reid M, et al. Real-time patient experience surveys of hospitalized medical patients. J Hosp Med. 2016; 11(4):251–256.

19. Hsieh HF, Shannon SE. Three approaches to qualitative content analysis. Qual Health Res. 2005; 15(9):1277–1288.

20. Merker VL, Dai A, Radtke HB, Knight P, Jordan JT, Plotkin SR. Increasing access to specialty care for rare diseases: a case study using a foundation sponsored clinic network for patients with neurofibromatosis 1, neurofibromatosis 2, and schwannomatosis. BMC Health Serv Res. 2018; 18(1):668.

21. Grol R. Successes and failures in the implementation of evidence-based guidelines for clinical practice. Med Care. 2001; 39(8 Suppl 2):II46–54.

22. Grimshaw JM, Thomas RE, MacLennan G, et al. Effectiveness and efficiency of guideline dissemination and implementation strategies. Health Technol Assess. 2004; 8(6):iii-iv, 1-72.

23. Montori VM, Brito JP, Murad MH. The optimal practice of evidence-based medicine: incorporating patient preferences in practice guidelines. JAMA. 2013; 310(23):2503–2504.

24. Brouwers MC, Kho ME, Browman GP, et al. AGREE II: advancing guideline development, reporting and evaluation in health care. CMAJ. 2010; 182(18):E839–842.

25. Lugtenberg M, Zegers-van Schaick JM, Westert GP, Burgers JS. Why don’t physicians adhere to guideline recommendations in practice? An analysis of barriers among Dutch general practitioners. Implement Sci. 2009; 4:54.

26. Pronovost PJ. Enhancing physicians’ use of clinical guidelines. JAMA. 2013; 310(23):2501–2502.

27. Kim C, Berta WB, R Gagliardi A. Exploring approaches to identify, incorporate and report patient preferences in clinical guidelines: Qualitative interviews with guideline developers. Patient Education and Counseling. 2020.

28. Seidlin M, Holzman R, Knight P, et al. Characterization and utilization of an international neurofibromatosis web-based, patient-entered registry: An observational study. PLoS One. 2017; 12(6):e0178639.

29. Gagliardi AR, Brouwers MC, Palda VA, Lemieux-Charles L, Grimshaw JM. How can we improve guideline use? A conceptual framework of implementability. Implementation Science. 2011; 6(1):26.

30. Legius E, Messiaen L, Wolkenstein P, et al. Revised diagnostic criteria for neurofibromatosis type 1 and Legius syndrome: an international consensus recommendation. Genetics in Medicine. 2021.

