## Supplemental Table 1 for "Awareness and Agreement with Neurofibromatosis Care Guidelines among Neurofibromatosis Specialists"

| Guideline | Number of Respondents | Percentage of Respondents who Selected “Strongly Agree” | 95% Confidence Interval* |
| --- | --- | --- | --- |
| **Neurofibromatosis 1 Guidelines** | | | |
| MRI preferred over CT scan | 59 | 83.1% | 73.5% - 92.6% |
| Blood pressure at least annually | 59 | 79.7% | 69.4% - 89.9% |
| Educate about MPNST | 59 | 76.3% | 65.4% - 87.1% |
| Development and school progress annually | 59 | 75.9% | 65.0% - 86.8% |
| Renovascular HTN screening | 57 | 73.7% | 62.3% - 85.1% |
| Height/weight annually | 59 | 70.9% | 59.3% - 82.5% |
| Annual eye testing <8yrs | 59 | 67.3% | 55.3% - 79.2% |
| Routine neurological exams annually | 59 | 65.5% | 53.3% - 77.6% |
| Pubertal development check-in annually | 59 | 64.8% | 52.6% - 77.0% |
| Skin exams annually | 59 | 64.4% | 52.2% - 76.6% |
| Eye exams Q6-12 month until 8 years old | 59 | 61.8% | 49.4% - 74.2% |
| Head circumference each visit til puberty | 59 | 52.7% | 40.0% -65.5% |
| Orthopedics referral for scoliosis | 59 | 52.5% | 39.8% -65.3% |
| NF clinic visit at least once per year | 59 | 52.5% | 39.8% -65.3% |
| Followed at specialized NF clinic | 59 | 52.5% | 39.8% -65.3% |
| Annual clinical check for scoliosis | 59 | 52.5% | 39.8% -65.3% |
| Screen adults for depression | 48 | 52.1% | 38.0% - 66.2% |
| Breast cancer screening ≥age 30 | 48 | 47.9% | 33.8% - 62.0% |
| Family planning annually | 59 | 47.5% | 34.7% - 60.2% |
| Vitamin D supplementation | 59 | 44.1% | 31.4% - 56.7% |
| Pheochromocytoma screening | 58 | 39.7% | 27.1% - 52.2% |
| Prefer MRA screen for renovascular HTN | 59 | 33.9% | 21.8% - 46.0% |
| High-risk obstetrician referral | 59 | 33.9% | 21.8% - 46.0% |
| Glomus tumor screening | 49 | 32.7% | 19.5% - 45.8% |
| WBMRI screen at 16-20 years old | 59 | 22.0% | 11.5% - 32.6% |
| Preanesthesia neuroimaging not needed | 59 | 16.9% | 7.4% - 26.5% |
| **Neurofibromatosis 2 Guidelines** | | | |
| Followed at specialized NF clinic | 51 | 72.5% | 60.3% - 84.8% |
| NF clinic visits at least once per year | 50 | 72.0% | 59.6% - 84.4% |
| Specialized ophthalmologist for children | 45 | 66.7% | 52.9% - 80.4% |
| Annual audiology with PTA and WRS | 51 | 64.7% | 51.6% - 77.8% |
| Genetic testing for children | 45 | 64.4% | 50.5% - 78.4% |
| Inform of need for lifelong imaging | 51 | 62.7% | 49.5% - 76.0% |
| Annual brain MRI; biannual if no tumors | 51 | 51.0% | 37.3% - 64.7% |
| Surveillance spine MRI Q24-36 months; can be reduced if no tumors | 51 | 47.1% | 33.4% - 60.8% |
| Surveillance spine MRI Q24-36 months beginning at age 10 | 45 | 37.8% | 23.6% - 51.9% |
| Spine MRI interval may be increased if no tumors | 45 | 35.6% | 21.6% - 49.5% |
| Annual brain MRI beginning at age 10; biannual if no tumors | 45 | 35.6% | 21.6% - 49.5% |
| **Schwannomatosis Guidelines** | | | |
| Genetic testing for children/young adults | 34 | 38.2% | 21.9% - 54.6% |
| Brain MRI Q24-36 month, beginning at age 10 for *SMARCB1* or age 15-19 for *LZTR1* | 33 | 30.3% | 14.6% - 46.0% |
| Spine MRI Q24-36 month, beginning at age 10 for *SMARCB1* or age 15-19 for *LZTR1* | 33 | 27.3% | 12.1% - 42.5% |

*Margin of error for the 95% confidence interval was calculated as

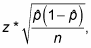

where z is the z-score, p̂ is the sample proportion, and n is the sample size.
